# Causes of variation in epigenetic aging across the lifespan

**DOI:** 10.1101/2020.05.10.20097030

**Authors:** Shuai Li, Tuong L Nguyen, Ee Ming Wong, Pierre-Antoine Dugué, Gillian S Dite, Nicola J Armstrong, Jeffrey M Craig, Karen A Mather, Perminder S Sachdev, Richard Saffery, Joohon Sung, Qihua Tan, Anbupalam Thalamuthu, Roger L Milne, Graham G Giles, Melissa C Southey, John L Hopper

## Abstract

**Background:** DNA methylation-based biological age (DNAm age) is potentially an important biomarker for adult health. Studies in specific age ranges have found widely varying results about its causes of variation. We investigated these causes across the lifespan.

**Methods:** We pooled genome-wide DNA methylation data for 4,217 people aged 0-92 years from 1,871 families. DNAm age was calculated using the Horvath epigenetic clock. We estimated familial correlations in DNAm age for monozygotic (MZ) twin, dizygotic (DZ) twin, sibling, parent-offspring, and spouse pairs by cohabitation status. Genetic and environmental variance component models were fitted and compared.

**Results:** Twin pair correlations were −0.12 to 0.18 around birth, not different from zero (all P>0.29). For all pairs of relatives, their correlations increased with time spent living together (all P<0.02) at different rates (MZ>DZ and siblings>parent-offspring; P<0.001) and decreased with time spent living apart (P=0.02) at similar rates. These correlation patterns were best explained by cohabitation-dependent shared environmental factors, the effects of which were 1.41 (95% confidence interval [CI], 1.16 to 1.66) times greater for MZ pairs than for DZ and sibling pairs, and the latter were 2.03 (95% CI, 1.13 to 9.47) times greater than for parent-offspring pairs. Genetic factors explained 13% (95% CI, −10% to 35%) of variation (P=0.27).

**Conclusion:** Variation in DNAm age is mostly caused by environmental factors, including those shared to different extents by relatives while living together and whose effects persist into old age. The equal environment assumption of the classic twin study might not hold for epigenetic aging.

---

Epigenetic alteration is considered to be a hallmark of aging^1^. Several measures of biological age based on DNA methylation (DNAm age) have been developed^2,3^, and found to be associated with mortality and disease risk in adulthood^2-6^. DNAm age, therefore, is potentially an important biomarker for adult health.

Lifestyle factors, disease risk factors, and genetic variants have been reported to be associated with DNAm age^2-4,7-10^. Pedigree-based and single nucleotide polymorphism (SNP)-based studies in specific age ranges, however, have given widely varying estimates of the percentage of variation in DNAm age caused by genetic factors, ranging from 0% to 100% depending on population, age range, and method^6-9,11-13^. There is also evidence that environmental factors shared within families explain a substantial proportion of variation^14^. Individual studies have typically focused on specific age ranges, so of themselves are not able to provide a comprehensive view of variation over the lifespan during which family members live together and apart at different ages, depending on their relationships.

We previously pooled DNA methylation data from a variety of twin and family studies in which participants were at different life stages, from birth to older age. We found evidence that variation in genome-wide average methylation is caused to a great extent by prenatal environmental factors, as well as by environmental factors shared by relatives (including spouse pairs) when they cohabit, and that these effects can persist at least to some extent across the whole lifetime^15^.

We have now applied the same approach to investigate the genetic, shared environmental, and individual-specific environmental causes of variation in DNAm age across the lifespan.

## Methods

### Study sample

We analyzed genome-wide DNA methylation data from 10 studies, some accessed through public repositories. The total sample included 4,217 people aged 0–92 years from 1,871 families, including monozygotic (MZ) twins, dizygotic (DZ) twins, siblings, parents, and spouses (Table 1). Most studies measured methylation using DNA extracted from peripheral blood and the HumanMethylation450 array (Supplementary Appendix).

**Table 1.** Sample characteristics by study.

| Study* | Biological sample | Microarray | Type of family members | N (females) | Mean chronological age (SD) | Mean DNAm age (SD) | Mean epigenetic age acceleration (SD) |
| --- | --- | --- | --- | --- | --- | --- | --- |
| PETS EPIC | Cord blood | EPIC | MZ | 46 (24) | 0 (0) | 1.1 (0.4) | -0.04 (0.38) |
|  |  |  | DZ | 44 (21) | 0 (0) | 1.2 (0.4) | 0.04 (0.43) |
| PETS 27K CMBCs | Cord blood mononuclear cells (CMBCs) | 27K | MZ | 34 (18) | 0 (0) | 0.1 (0.3) | -0.09 (0.25) |
|  |  |  | DZ | 18 (4) | 0 (0) | 0.3 (0.5) | 0.16 (0.50) |
| PETS 27K HUVECs | Human umbilical vascular endothelial cells (HUVECs) | 27K | MZ | 26 (14) | 0 (0) | 6.2 (4.1) | -0.95 (4.10) |
|  |  |  | DZ | 16 (4) | 0 (0) | 8.6 (5.1) | 1.54 (5.07) |
| PETS 27K placenta | Placenta | 27K | MZ | 16 (12) | 0 (0) | 0.0 (0.2) | 0.05 (0.21) |
|  |  |  | DZ | 12 (5) | 0 (0) | -0.1 (0.2) | -0.07 (0.21) |
| PETS 450K birth | Buccal cells | 450K | MZ | 18 (8) | 0 (0) | 0.8 (0.4) | 0.02 (0.37) |
|  |  |  | DZ | 10 (4) | 0 (0) | 0.8 (0.2) | -0.04 (0.15) |
| PETS 450K 18 months | Buccal cells | 450K | MZ | 12 (6) | 1.5 (0) | 2.2 (0.7) | 0.10 (0.66) |
|  |  |  | DZ | 8 (2) | 1.5 (0) | 2.0 (0.5) | -0.15 (0.46) |
| BSGS | Peripheral blood | 450K | MZ | 134 (62) | 13.8 (1.9) | 18.4 (3.6) | -0.34 (2.68) |
|  |  |  | DZ | 222 (107) | 13.2 (2.0) | 18.0 (3.7) | 0.09 (2.63) |
|  |  |  | Sibling | 119 (59) | 15.5 (2.8) | 21.2 (4.8) | 0.22 (2.84) |
|  |  |  | Spouse/parents | 139 (73) | 46.6 (5.6) | 50.8 (5.2) | 0.00 (3.52) |
| E-Risk | Peripheral blood | 450K | MZ | 852 (414) | 18 (0) | 24.1 (3.7) | -0.01 (3.67) |
|  |  |  | DZ | 612 (300) | 18 (0) | 24.1 (4.0) | 0.03 (4.02) |
| DTR younger adults | Peripheral blood | 450K | MZ | 146 (66) | 33.1 (2.0) | 32.6 (4.8) | 0.00 (4.31) |
| AMDTSS | Peripheral blood | 450K | MZ | 132 (132) | 55.6 (8.4) | 54.8 (6.7) | -0.34 (4.58) |
|  |  |  | DZ | 132 (132) | 57.0 (7.2) | 56.5 (6.0) | 0.60 (5.09) |
|  |  |  | Sibling | 215 (215) | 56.6 (8.0) | 55.5 (6.5) | -0.16 (4.92) |
| TwinsUK | Peripheral blood | 27K | MZ | 66 (66) | 58.4 (9.1) | 56.0 (8.9) | -0.75 (5.10) |
|  |  |  | DZ | 86 (86) | 56.6 (7.7) | 55.6 (8.1) | 0.57 (4.14) |
| MuTHER | Adipose tissue | 450K | MZ | 186 (186) | 61.0 (9.3) | 58.9 (6.1) | -0.19 (3.67) |
|  |  |  | DZ | 306 (306) | 57.4 (9.3) | 57.3 (6.2) | 0.12 (3.57) |
| DTR older adults | Peripheral blood | 450K | MZ | 154 (78) | 63.2 (4.1) | 60.5 (6.7) | 0.00 (5.63) |
| OATS | Peripheral blood | 450K | MZ | 216 (136) | 71.2 (6.0) | 65.5 (6.6) | 0.00 (5.36) |
| LSADT 1997 | Peripheral blood | 450K | MZ | 36 (22) | 76.3 (2.0) | 71.8 (3.7) | -0.25 (3.86) |
|  |  |  | DZ | 50 (40) | 76.2 (1.6) | 72.2 (5.7) | 0.18 (5.50) |
| LSADT 2007 | Peripheral blood | 450K | MZ | 36 (22) | 86.2 (2.0) | 79.5 (4.9) | -0.24 (4.85) |
|  |  |  | DZ | 50 (40) | 86.1 (1.6) | 80.0 (5.4) | 0.18 (5.47) |
| MCCS | Peripheral blood | 450K | Spouse | 124 (62) | 60.1 (6.2) | 59.9 (8.1) | 0.00 (6.19) |

Abbreviations – EPIC: the HumanMethylationEPIC array; 27K: the HumanMethylation27 array; 450K: the HumanMethylation450 array; MZ: monozygotic twin;
DZ: dizygotic twin; N: sample size; SD: standard deviation
\*Studies – PETS: Peri/postnatal Epigenetic Twins Study, including three datasets measured using the 27K array (using three biological samples), 450K array (at two points: at birth and age 18 months), and EPIC array, respectively; BSGS: Brisbane System Genetics Study; E-Risk: Environmental Risk Longitudinal Twin Study; DTR: Danish Twin Registry, in two groups: younger and older adults; AMDTSS: Australian Mammographic Density Twins and Sisters Study; MuTHER: Multiple Tissue Human Expression Resource Study; OATS: Older Australian Twins Study; LSDAT: Longitudinal Study of Aging Danish Twins, with samples collected at years 1997 and 2007, respectively; MCCS: Melbourne Collaborative Cohort Study

### DNAm age and epigenetic age acceleration

We used the Horvath epigenetic clock^12^ to determine DNAm age (https://dnamage.genetics.ucla.edu/new) because it was developed across tissues and ages, and the 353 methylation sites used by this clock are common to the three methylation arrays used by the 10 studies (Table 1). We chose the ‘Normalize Data’ option of the online calculator to normalize each dataset to be comparable to the training data of this epigenetic clock. Epigenetic age acceleration was defined as DNAm age adjusted for the effects of chronological age. Sensitivity analyses were performed using only those studies in which DNA methylation was measured in blood to examine the robustness of results to cell composition (see Supplementary Appendix).

### Statistical analysis

Residuals of epigenetic age acceleration adjusted for sex were used in subsequent analyses. We used a multivariate normal model for pedigree analysis^17,18^ and the program FISHER^19^ to estimate correlations and to fit variance components models; see below. The likelihood ratio test was used to compare nested models. All P-values were two-sided and P<0.05 was considered significant.

According to the pattern in familial correlations by chronological age, and following previous theoretical and empirical studies^15,17,20^, the familial correlations for different types of pairs (MZ, DZ, sibling, parent-offspring and spouse) across the lifespan were modelled as a function of cohabitation status (see Supplementary Appendix).

We then assumed that the variation of DNAm age can be caused by combinations of additive genetic factors (A), shared environmental factors (C), and individual-specific environmental factors (E). We assessed model fits using the Akaike Information Criterion for the following models and assumptions (see Supplementary Appendix):

1. AE model: variation is caused by only A and E, and the effects of A are constant across the lifespan;
2. Cohabitation-dependent AE model: variation is caused only by A and E, and the effects of A depend on cohabitation;
3. Cohabitation-dependent ACE model: variation is caused by A, C and E, and the effects of A and C both depend on cohabitation;
4. Cohabitation-dependent CE model: variation is caused by only C and E, and the effects of C depend on cohabitation.

## Results

### Sample characteristics

DNAm age was moderately to strongly correlated with chronological age within each dataset, with correlations ranging from 0.44 to 0.84 (Figure S1). The variance of DNAm age increased with chronological age, being small for newborns, greater for adolescents, and relatively constant with age for adults (Figure S2).

### Within-study familial correlations

Table 2 shows the within-study familial correlation estimates. There was no difference in the correlation between MZ and DZ pairs for newborns or adults, but there was a difference (P<0.001) for adolescents (Table 2): 0.69 (95% confidence interval [CI], 0.63 to 0.74) for MZ pairs, and 0.35 (95% CI, 0.20 to 0.48) for DZ pairs. For MZ and DZ pairs combined, there was consistent evidence across datasets and tissues that the correlation was around −0.12 to 0.18 at birth and 18 months, not different from zero (all P>0.29), and about 0.3 to 0.5 for adults (different from zero in seven of eight datasets; all P<0.01). Across all datasets, the results suggested that twin pair correlations increased with age from birth up until adulthood and were maintained to older age.

**Table 2.** Within-study familial correlations in DNAm age.

| Study* | Type of pairs | Number of pairs | Mean age | Correlation (95% CI) | P | P for MZ vs DZ |
| --- | --- | --- | --- | --- | --- | --- |
| PETS EPIC | MZ | 23 | 0 | 0.18 (-0.28 to 0.57) | 0.45 | 0.57 |
|  | DZ | 22 | 0 | 0.01 (-0.35 to 0.36) | 0.98 |  |
|  | MZ and DZ | 45 | 0 | 0.15 (-0.22 to 0.34) | 0.64 |  |
| PETS 27K | MZ | 22 | 0 | 0.35 (-0.04 to 0.64) | 0.85 | 0.69 |
|  | DZ | 11 | 0 | 0.00 (-0.46 to 0.46) | 0.09 |  |
|  | MZ and DZ | 33 | 0 | 0.18 (-0.15 to 0.47) | 0.76 |  |
| PETS 450K birth | MZ | 9 | 0 | -0.05 (-0.51 to 0.43) | 0.09 | 0.29 |
|  | DZ | 5 | 0 | 0.62 (0.01 to 0.89) | 0.99 |  |
|  | MZ and DZ | 14 | 0 | -0.08 (-0.54 to 0.41) | 0.29 |  |
| PETS 450K 18 months | MZ | 6 | 1.5 | -0.06 (-0.62 to 0.55) | 0.87 | 0.58 |
|  | DZ | 4 | 1.5 | -0.52 (-0.91 to 0.40) | 0.31 |  |
|  | MZ and DZ | 10 | 1.5 | -0.12 (-0.62 to 0.46) | 0.71 |  |
| BSGS | MZ | 67 | 13.8 | 0.69 (0.63 to 0.74) | <0.001 | <0.001 |
|  | DZ | 111 | 13.2 | 0.35 (0.20 to 0.48) | <0.001 |  |
|  | Siblings | 260 | 14.0 | 0.32 (0.20 to 0.42) | <0.001 |  |
|  | Parent-offspring | 363 | 13.4 | 0.15 (0.02 to 0.27) | 0.02 |  |
|  | Spouses | 59 | 46.6 | -0.01 (-0.25 to 0.24) | 0.96 |  |
| E-Risk | MZ | 426 | 18.0 | 0.46 (0.40 to 0.52) | <0.001 | 0.28 |
|  | DZ | 306 | 18.0 | 0.40 (0.33 to 0.47) | <0.001 |  |
|  | MZ and DZ | 732 | 18.0 | 0.43 (0.39 to 0.48) | <0.001 |  |
| DTR younger adults | MZ | 73 | 33.1 | 0.62 (0.52 to 0.70) | <0.001 | — |
| AMDTSS | MZ | 66 | 55.6 | 0.43 (0.26 to 0.58) | <0.001 | 0.20 |
|  | DZ | 66 | 57.0 | 0.24 (0.04 to 0.42) | 0.02 |  |
|  | MZ and DZ | 132 | 56.3 | 0.32 (0.18 to 0.45) | <0.001 |  |
|  | Siblings | 552 | 56.4 | 0.12 (0.02 to 0.22) | 0.02 |  |
| TwinsUK | MZ | 33 | 55.3 | 0.23 (-0.03 to 0.46) | 0.09 | 0.41 |
|  | DZ | 43 | 59.2 | 0.04 (-0.33 to 0.39) | 0.85 |  |
|  | MZ and DZ | 76 | 57.5 | 0.12 (-0.07 to 0.37) | 0.17 |  |
| MuTHER | MZ | 93 | 58.4 | 0.54 (0.44 to 0.63) | <0.001 | 0.08 |
|  | DZ | 153 | 56.6 | 0.37 (0.25 to 0.48) | 5.8E-08 |  |
|  | MZ and DZ | 246 | 57.3 | 0.44 (0.36 to 0.52) | <0.001 |  |
| DTR older adults | MZ | 77 | 63.2 | 0.55 (0.43 to 0.65) | <0.001 | – |
| OATS | MZ | 108 | 71.2 | 0.40 (0.26 to 0.53) | <0.001 | – |
| LSADT 1997 | MZ | 18 | 76.3 | 0.04 (-0.66 to 0.70) | 0.93 | 0.36 |
|  | DZ | 25 | 76.2 | 0.39 (0.14 to 0.60) | <0.001 |  |
|  | MZ and DZ | 43 | 76.2 | 0.34 (0.09 to 0.55) | 0.01 |  |
| LSADT 2007 | MZ | 18 | 86.2 | 0.41 (0.05 to 0.68) | 0.04 | 0.95 |
|  | DZ | 25 | 86.1 | 0.40 (0.11 to 0.62) | 0.01 |  |
|  | MZ and DZ | 43 | 86.1 | 0.40 (0.17 to 0.59) | <0.001 |  |
| MCCS | Spouses | 62 | 60.1 | 0.12 (-0.12 to 0.35) | 0.33 | – |

Abbreviations – MZ: monozygotic twin; DZ: dizygotic twin; CI: confidence interval
\*Studies – PETS: Peri/postnatal Epigenetic Twins Study, including three datasets measured using the 27K array, 450K array (at two points: at birth and age 18 months), and EPIC array, respectively; BSGS: Brisbane System Genetics Study; E-Risk: Environmental Risk Longitudinal Twin Study; DTR: Danish Twin Registry, in two groups: younger and older adults; AMDTSS: Australian Mammographic Density Twins and Sisters Study; MuTHER: Multiple Tissue Human Expression Resource Study; OATS: Older Australian Twins Study; LSDAT: Longitudinal Study of Aging Danish Twins, with samples collected at years 1997 and 2007, respectively; MCCS: Melbourne Collaborative Cohort Study

The correlation for adolescent sibling pairs was 0.32 (95% CI, 0.20 to 0.42), and not different from that for adolescent DZ pairs (P=0.89), but less than that for adolescent MZ pairs (P<0.001). Middle-aged sibling pairs were correlated at 0.12 (95% CI, 0.02 to 0.22), less than that for adolescent sibling pairs (P=0.02). Parent-offspring pairs were correlated at 0.15 (95% CI, 0.02 to 0.27), less than that for pairs of other types of 1^st^-degree relatives in the same study, e.g., DZ pairs and sibling pairs (both P<0.04). The spouse pair correlations were −0.01 (95% CI, −0.25 to 0.24) and 0.12 (95% CI, −0.12 to 0.35).

From the sensitivity analysis, the familial correlation results were robust to the adjustment for blood cell composition (Table S1).

### Familial correlations across the lifespan

From modelling the familial correlations for the different types of pairs as a function of their cohabitation status (Table S2), the estimates of θ (see Supplementary Appendix for the definition) ranged from 0.76 to 1.20 across pairs, none different from 1 (all P>0.1). We therefore fitted a model with θ=1 for all pairs; the fit was not different from the model above (P=0.69). Under the latter model, the familial correlations increased with time living together at different rates (P<0.001) across pairs. The decreasing rates did not differ across pairs (P=0.27). The correlations for DZ and sibling pairs were similar (P=0.13), and when combined their correlation was different from that for parent-sibling pairs (P=0.002) even though these pairs are all genetically 1^st^-degree relatives, and was smaller than that for the MZ pairs (P=0.001).

We then fitted a model in which DZ and sibling pairs were combined and the decreasing rates were the same across all pairs. The goodness-of-fit of this model was not inferior to that of the model above (P=0.14) and the model included fewer parameters. Under this model, the familial correlations for MZ, DZ and sibling, and parent-offspring pairs all increased with time living together (all P<0.02) with different increasing rates (P<0.001); most rapidly for MZ pairs (*λ=* 0.041, 95% CI, 0.035 to 0.048), less rapidly for DZ and sibling pairs (*λ*= 0.026, 95% CI, 0.020 to 0.031), and least rapidly for parent-offspring pairs (*λ*=0.011, 95% CI, 0.002 to 0.0021), and decreased with time living apart (P=0.02); see Figure 1.

**Figure 1.**
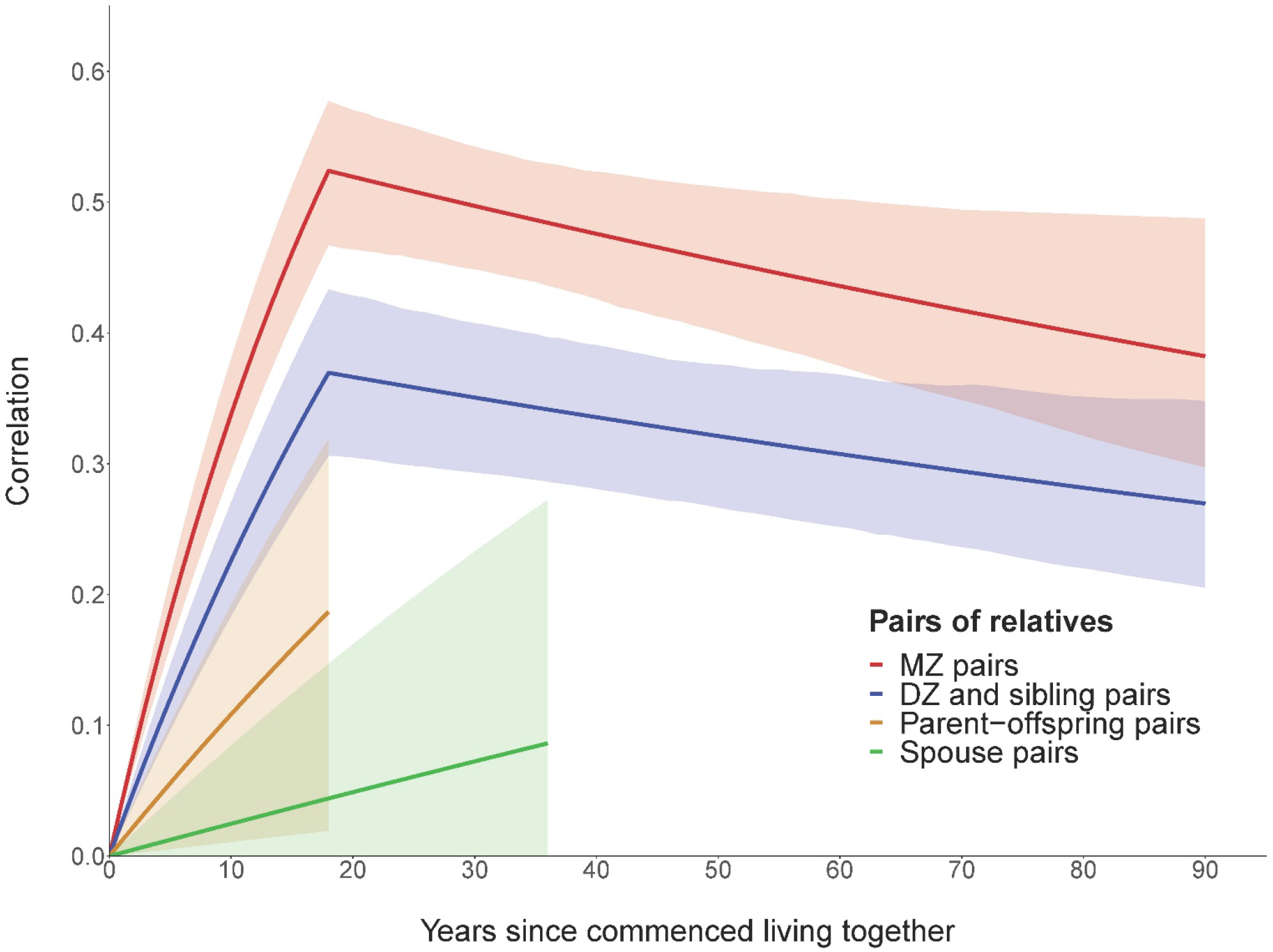
Familial correlations in DNAm age for the different types of pairs across the lifespan. Lines are the correlation estimates, and shadows are the corresponding 95% confidence intervals. MZ: monozygotic twin; DZ: Dizygotic twin.

### Causes of variation across the lifespan

Results from modelling the causes of variation across the lifespan are shown in Figure 2 and Table S3. Under the AE model, additive genetic factors explained 52% (95% CI, 48% to 53%) of variation. This, however, was the worst fitting model.

**Figure 2.**
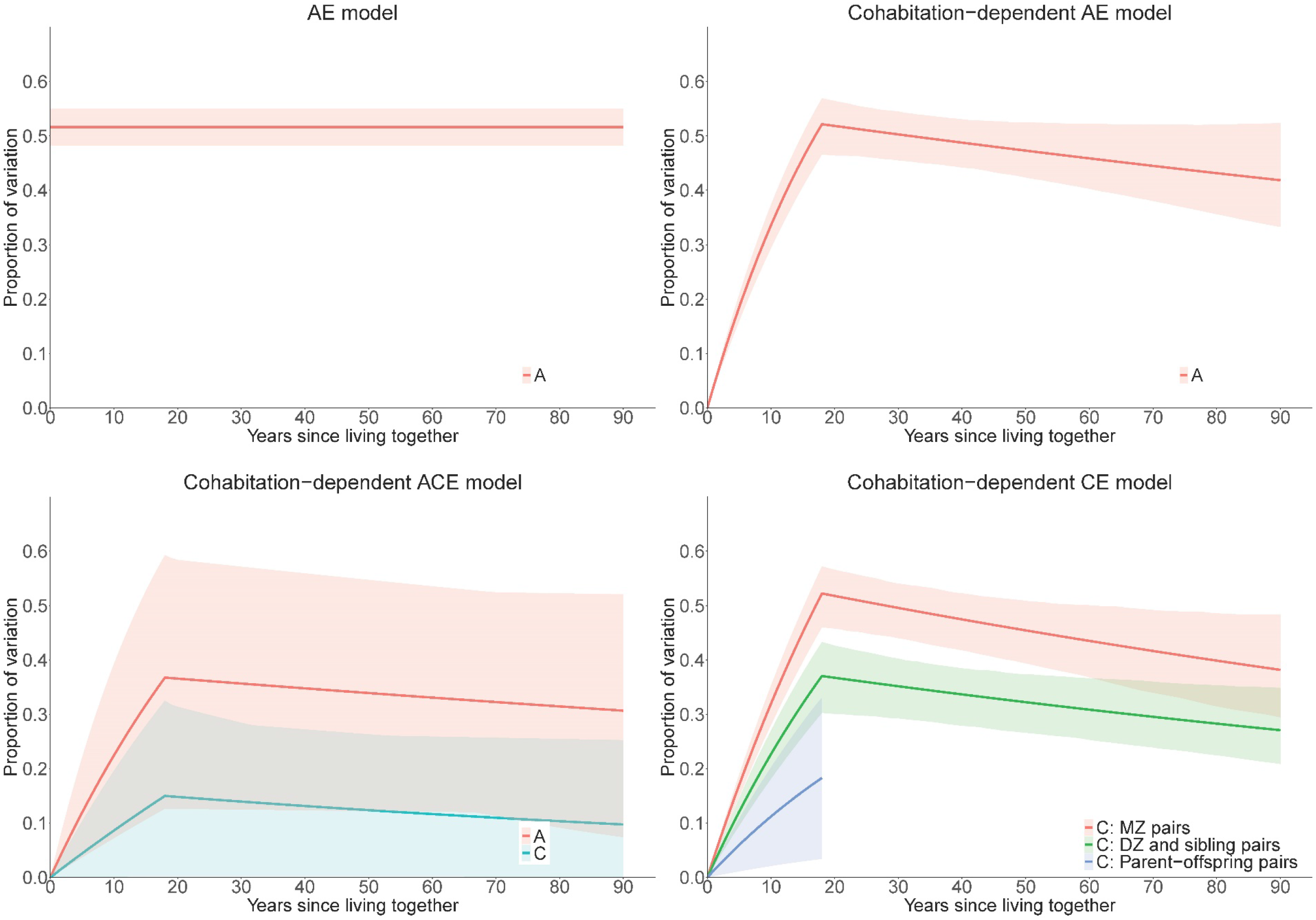
Proportion of variation in DNAm age across the lifespan explained by genetic and environmental factors under different models. Lines are the correlation estimates, and shadows are the corresponding 95% confidence intervals. A: additive genetic factors; C: shared environmental factors; E: individual-specific environmental factors; MZ: monozygotic twin; DZ: Dizygotic twin. Model details - AE model: variation was assumed to be caused by only A and E, and the effects of A are constant across the lifespan; Cohabitation-dependent AE model: variation was assumed to be caused by only A and E, and the effects of A depend on cohabitation; Cohabitation-dependent ACE model: variation was assumed to be caused by A, C and E, and the effects of A and C both depend on cohabitation; Cohabitation-dependent CE model: variation is caused by only C and E, and the effects of C depend on cohabitation.

Under the cohabitation-dependent AE model, the effects of genetic factors increased with time living together and decreased with time living apart, and explained minimal variation around birth, ~40% of variation in adolescence and adulthood, and ~50% of variation at age of 18 years.

Under the cohabitation-dependent ACE model, both the effects of genetic factors and the effects of shared environmental factors increased with time living together but did not change with time living apart. The goodness-of-fits of the cohabitation-dependent AE and cohabitation-dependent ACE models were similar.

The best fitting model was the cohabitation-dependent CE model. Under this model, different pairs shared the effects of environmental factors to different extents. The effects for MZ pairs were 1.41 (95% CI, 1.16 to 1.66) times those for DZ and sibling pairs, and the latter were 2.03 (95% CI, 1.13 to 9.47) times those for parent-offspring pairs. For all pairs, the proportion of variation explained by the shared environmental factors increased with time living together (P<0.001) and decreased at a slower rate with time living apart (P=0.02).

Under the above cohabitation-dependent CE model, we further assumed that the variation is additionally caused by genetic factors whose effects are constant across the lifespan (see Supplementary Appendix). Genetic factors were estimated to explain 13% (95% CI, −10% to 35%) of the variation (P=0.27). That is, after taking into account the existence of non-genetic cohabitation-dependent effects, there was no evidence for a substantive role of genetic factors.

## Discussion

Our study provides novel insights into the causes of variation in DNAm age across the lifespan, which appear to be largely due to environmental (i.e. non-genetic) factors. These include cohabitation-related environmental factors that are evident prior to adulthood, and whose effects persist across the whole of the lifespan. Two longitudinal studies have also found that DNAm age is largely set before adulthood^21^.

Our data suggest that people in the same family are not correlated in DNAm age when they start cohabiting; the longer they live together the more similar they become but at a rate that differs substantially depending on their relationship. This is likely due to the different types of relatives sharing environmental factors relevant to DNAm age to different degrees. When pairs of relatives live apart, they no longer share the cohabitation environment, and this is reflected by a slow dissipation of the effects of shared environmental factors across adulthood at a rate that appears to be similar for all pairs.

For MZ pairs, some DNA methylation measures have been found to be similar at birth but divergent over the lifetime, a phenomenon called ‘epigenetic drift’^15,22^. DNAm age, however, shows a different pattern; MZ pairs are not similar at birth (and neither are DZ pairs) but become more similar the longer they live together, and do so more rapidly than do DZ or other pairs of relatives. MZ pairs then appear to slowly become less similar in DNAm age the longer they live apart in adulthood, at the same rate as for other pairs of relatives, but still maintain a substantial similarity even into late life. These observations suggest that DNAm age reflects biological aging processes beyond what is reflected by DNA methylation alone.

Our finding that environmental factors shared while cohabiting play a major role in determining the variation in DNAm age is also supported by the observation that the variance of DNAm age increased dramatically with age prior to adulthood, and was relatively stable across adulthood (Figure S2). The latter has also been found by previous studies^21^.

Given DNAm age has been found to be associated with the risks of death and various diseases in adulthood, identifying the environmental factors affecting DNAm age prior to adulthood might give novel insights into which, and how, early-life factors impact late-life health outcomes. This would have obvious implications for prevention and its timing. There is some evidence that DNAm age is associated with physical developmental characteristics, and exposures to stress and violence for children, although most studies had a moderate sample size^23-27^.

The classic twin design assumes that MZ and DZ pairs share environmental effects relevant to the trait of interest to exactly the same extent; i.e., the equal environment assumption. Our study shows that this assumption might not hold for DNAm age because there was strong evidence that MZ and DZ pairs share their pre-adult environmental effects to different extents. Furthermore, DZ and sibling pairs were more correlated than parent-offspring pairs, despite all being genetically 1^st^-degree relatives of one another. This is not consistent with the correlations predicted by additive genetic factors. Given there is little evidence of genetic effects, our results are not consistent with gene-environment interaction either^28^; we found that models including genetic effects, no matter whether as constant or cohabitation-dependent, were less consistent with the data compared with the cohabitation-CE model.

Previous twin and pedigree studies have assumed the equal environment assumption holds perfectly, and consequently reported the heritability of DNAm age to be ~40% in adolescence and middle-age^6,9,12^. Note that, under our cohabitation-dependent AE model (which makes the equal environment assumption) genetic factors would explain ~40% of variation in adolescence and middle-age. This model, however, was not a good fit and was rejected in favor of models that included cohabitation-dependent environmental effects.

Studies have predicted that measured SNPs could explain 0–70% of variation in DNAm age measured from whole blood and brain tissue^7-9,11^. Those analyses explicitly assumed, however, that all of the phenotypic covariance is due to genetic factors. In particular, one study predicted the SNP-based heritability of DNAm age based on mothers and children increased with the children’s age, being zero when the children were around birth and 37% when the children were 15 years old^7^ – in line with our data and the estimates under the cohabitation-dependent AE model that was rejected. Without relying on the equal environment assumption, we found that genetic factors explained at most a small, and not statistically significant, proportion (~10%) of variation. Therefore, studies using the equal environment assumption might have overestimated the influence of genetic factors on DNAm age variation.

We used the epigenetic clock to determine DNAm age, as this clock is mostly applicable to our multi-tissue methylation data and study sample including newborns, children and adults. Other DNAm age measures^29-31^ were developed using blood methylation data for middle-aged people only.

Our findings should be interpreted with caution given that they are from statistical modelling which alone cannot prove that a consistent model is a true representation of nature. All that can be said is whether or not the data “are consistent with” a particular explanation. Nonetheless, statistical modelling is an attempt to identify the plausible and implausible explanations of data, and our results suggest that cohabitation environmental factors being shared by pairs of relatives to different extents is more plausible than other explanations.

Our study has some potential limitations given we pooled data across studies. To minimize heterogeneity, we normalized methylation data from each study to be comparable to the training data of the epigenetic clock and used study-specific variances. Some studies had moderate sample sizes, so that failure to detect differences between some familial correlations at certain ages might be due to a lack of statistical power. Similarly, we cannot categorically exclude a role for genetic factors.

In conclusion, the variation in epigenetic aging as measured by the epigenetic clock that we observed is most consistent with having been caused, at least to a large extent, by environmental factors, including those shared to different extents by relatives while living together. The effects of the cohabitation environment increase with the time living together, and persist into old age. The equal environment assumption of the classic twin study might not hold for epigenetic aging. Given the observed relationships between DNAm age and health outcomes, these findings highlight the importance and potential of pre-adulthood prevention related to environmental factors for adult diseases.

## Data Availability

The following datasets are accessed from Gene Expression Omnibus: PETS 27K (GSE36642), PETS 450K (GSE42700), BSGS (GSE56105), E-Risk (GSE105018), DTR (GSE61496), TwinsUK (GSE58045) and LSDAT (GSE73115). The MuTHER dataset was assessed from ArrayExpress with the number E-MTAB-1866.

## Acknowledgements

We thank the participants of each studies. We thank the researchers who make these datasets available from public repositories: PETS 27K (GSE36642), PETS 450K (GSE42700), BSGS (GSE56105), E-Risk (GSE105018), DTR (GSE61496), TwinsUK (GSE58045), MuTHER (E-MTAB-1866) and LSDAT (GSE73115). We thank Dr Allan McRae from The University of Queensland for providing additional data of the BSGS.

## Author contributions

Study conception and design: SL & JLH. Data analysis: SL. Manuscript drafting: SL & JLH. Data collection: PETS – SL, EMW, JMC, RS, MCS & JLH, MCCS – EMW, PAD, GGG, MCS, RLM & JLH, AMDTSS – SL, EMW, TLN, GSD, GGG, MCS & JLH, OATS – NJA, KAM, PSS & AT, DTR & LSDAT – QT. All authors have contributed to manuscript editing and data interpretation.

